# Psychological Distress during the COVID-19 pandemic in France: a national assessment of at-risk populations

**DOI:** 10.1101/2020.05.10.20093161

**Authors:** Benjamin Chaix, Guillaume Delamon, Arthur Guillemassé, Benoît Brouard, Jean-Emmanuel Bibault

## Abstract

**Background:** More than 2.5 billion people in the world are currently in lockdowns to limit the spread of the novel coronavirus disease 2019 (COVID-19). Psychological Distress (PD) and Post-Traumatic Stress Disorder have been reported after traumatic events, but the specific effect of pandemics is not well known.

**Objectives:** The aim of this study was to assess PD in France, a country where COVID-19 had such a dramatic impact that it required a country-wide lockdown.

**Methods:** This study was a survey conducted in France between 31 March 2020 and 7 April 2020. We recruited patients in 4 groups of chatbot users followed for breast cancer, asthma, depression and migraine. We used the Psychological Distress Index (PDI), a validated scale to measure PD during traumatic events, and correlated PD risk with patients’ characteristics in order to better identify the one who were the most at-risk.

**Results:** The study included 1771 participants. 91.25% (1616) were female with a mean age of 32.8 years (SD=13,71), 7.96% (141) were male with a mean age of 28.0 years (SD=8,14). In total, 38.06% (674) of the respondents had psychological distress (PDI ≥15). An ANOVA analysis showed that sex (p=0.00132), unemployment (p=7.16×10-6) and depression (p=7.49×10-7) were significantly associated with a higher PDI score. Patients using their smartphone or computer more than one hour a day also had a higher PDI score (p=0.02588).

**Conclusion:** Prevalence of PD in at-risk patients is high. These patients are also at increased risk to develop Post-Traumatic Stress Disorder. Specific steps should be implemented to monitor and prevent PD through dedicated mental health policies if we want to limit the public health impact of COVID-19 in time.

## 1. Introduction

The novel coronavirus disease 2019 (COVID-19) pandemic called for unprecedented policies by governments around the world to counter its spread. Most European countries have implemented social distancing and shelter in place measures. These measures are comparable to generalized quarantine and prevent the spread of the virus by restricting the movement and social interactions of people who are potentially exposed (1). As of May, 3rd, 2.5 billion people are in lockdowns (2). In France, these measures have been in force since 17 March 2020. Several studies have reported the negative effects of quarantine on stress or depression (3–5).

Peritraumatic distress (PD) is defined as the emotional and physiological distress experienced during and/or immediately after a traumatic event. It is associated with a higher risk and severity of Post-Traumatic Stress Disorder (PTSD) (6,7). The Peritraumatic Distress Inventory (PDI) was created to assess the emotional and physiological experience of individuals during a traumatic event (8). Studies have shown that PDI has a good internal consistency, stability, and validity. PDI items can be grouped into factors that better reflect and predict PD and PTSD: negative emotions (items 1, 2, 3, 5, 6, 8, and 10) and perceived life threat and bodily arousal (items 4, 7, 9, 11, 12, and 13) (9). It has also been shown that a PDI score equal or over 14 was predicting full or partial PTSD six-weeks post-injury (10).

The main objective of this study was to assess the effect of the Covid-19 crisis on psychological distress in at-risk patients. The secondary objectives were to describe the patients characteristics that can be used to predict the risk of PD and PTSD. In order to do so, we built an e-cohort consisting of users of 4 medical chatbots designed to support patients with (1) asthma, (2) breast cancer, (3) depression and (4) migraine. A chatbot is a software leveraging artificial intelligence to provide a natural language conversation with a user. They can be use to monitor patients during treatment or to collect patient-reported outcomes (11). Vik chatbots, developed by Wefight, have shown their interest in patient support and adherence to treatment (12). They are also able to provide medical information to breast cancer patients with a level of quality comparable to physicians, as shown in the phase 3 randomized controlled trial INCASE (NCT03556813) (13). The “Vik Asthm” chatbot is dedicated to information and management of asthma-related symptoms, the “Vik Breast” chatbot is specialized in the management of breast cancer patients, the “Vik Depression” chatbot accompanies patients with symptoms of depression and the “Vik Migraine” chatbot is helping patients with chronic migraine.

People with asthma are populations at increased risk of severe viral respiratory infections that can also induce exacerbations. The SARS-CoV2 can induce asthma exacerbations which are a source of additional stress for asthma patients. Initial data shows that asthma patients do not appear to be overrepresented in patients with Covid-19 (14,15). In order to mitigate the lack of pathology control and treatment adherence during travel restrictions, the French government has implemented solutions to facilitate the renewal of treatment in the long term (16). In addition, 60,000 hospitalizations are attributable to asthma every year in France (17). This pandemic also represents a significant concern to cancer patients, who are at high risk of complications due to several predisposing factors (18-20). In patients with breast cancer, management must be tailored and cannot be delayed. European countries have increased the use of telehealth systems to reduce the number of hospital visits. In Italy, these changes in care lead to many questions for patients, which can generate severe stress or anxiety (21). The first studies conducted in China following the coronavirus pandemic have shown the impact on the mental health of healthcare workers, with an increased risk of depression and anxiety (22). Patients already diagnosed with depressive disorder could be at a high risk of distress during the Covid-19 pandemic. Finally, migraine is a pathology with a high prevalence: it is estimated to be between 17% and 21% in adults aged 18 to 65 (23) with a sex ratio of 3 women to 1 man (24). Despite its high prevalence, migraine remains an under-diagnosed and under-treated condition in the general population. Migraine can have a significant impact on the patient’s quality of life (25). Migraines can worsen in times of stress. This period of pandemic can generate a new source of stress and aggravate the pathology.

## 2. Patients and methods

### 2.1 Participants

The study was conducted in France between 31 March 2020 and 7 April 2020. The participants were users of the 4 different Vik chatbots. They were contacted online to participate in a survey assessing their level of stress during the Covid-19 crisis. The inclusion criteria were to be of legal age and to have breast cancer, asthma, migraine or depression. The non-inclusion criteria concerned users who were unable to formulate their non-opposition, who had insulted the chatbot or who had dialogues that made no sense.

### 2.2 Intervention

A self-report questionnaire, the Peritraumatic Distress Index (PDI), was used. Peritraumatic distress is defined as the emotional and physiological distress experienced during and/or immediately after a traumatic event (8,9). It is the standard tool designed to assess psychological distress in times of crisis. It consists of 13 questions rated from 0 (not at all true) to 4 (extremely true). It explores the frequency of anxiety, depression, specific phobias, cognitive changes, avoidance and compulsive behaviors, physical symptoms and loss of social interaction in the past week. The total score, ranging from 0 to 52, is the sum of all items. A score equal of over 15 indicates significant distress. The French validation of the PDI has a good internal cohesion, with a Cronbach’s alpha of 0.83 (26).

### 2.3 Data collection

The PDI questionnaire was presented to the participants by text messages. Users were asked to click on a button corresponding to the score they wished to give their status. There was no actual conversation per question, nor was there a need for natural language processing for each question. Classical demographic information (age, sex, city, professional profile), level of knowledge and use of internet tools and the presence or absence of symptoms related to COVID-19 were also assessed.

### 2.4 Ethical and regulatory issues

Participants were not paid. The collected data were anonymized and then hosted by Wefight on a server that meets the requirements for storing health data. Consent was collected online before the start of the study. This study was registered in the ClinicalTrials.gov database (NCT04337047) and was approved by our internal review board. The need for ethical approval was waived for this non-interventional study. In accordance with French and European laws on information technology and liberties (Commission Nationale Informatique et Libertés, registration n° 2217452, General Regulations for Data Protection), users had the right to access the data to verify its accuracy and, if necessary, to correct, complete and update it. They also had a right to object to their use and a right to delete such data. The general conditions for the use of the data were presented and explained very clearly. They had to be accepted before accessing the questionnaire.

### 2.5 Statistical analysis

The description of the populations included was carried out by the classic elements of the calculation of mean, standard deviation, median and quartiles for quantitative variables, numbers and percentages and 95% confidence intervals for qualitative variables. The population density was defined by French Government’s DEPP (Direction of evaluation, prospection and performance) (27).

ANOVA was performed to detect patients’ attributes with a significant effect on PDI. In addition, a binomial logistic regression analyze was carried out to determine the patients features associated with a PDI>14, because this subpopulation is at a higher risk of partial of full PTSD six weeks after the traumatic event.

PDI items were grouped into two factors that have been shown to better reflect and predict PD and PTSD: negative emotions (items 1, 2, 3, 5, 6, 8, and 10) and perceived life threat and bodily arousal (items 4, 7, 9, 11, 12, and 13). For both groups an ANOVA was performed.

The Pearson correlation coefficient was calculated between the average PDI and the number of infected people in each French region and was tested to be equal to 0.

## 3. Results

### 3.1 Cohort description

The study included 1892 participants. We excluded 121 of them because they were not eligible (incomplete questionnaires and age requirements). The total sample size was 1771. Overall, 91.25% (1616) were female with a mean age of 32.8 years (SD=13,71), 7.96% (141) were male with a mean age of 28.0 years (SD=8,14) and 0.79% (14) were “other” with a mean age of 25.6 years (SD=8,85) (Table 1). In total, 3.3% (58) of participants were using a smartphone or computer less than an hour a day, 55.5% (983) for more than 1 hour but less than 6 hours a day and 41.22% (730) more than 6 hours a day. They were 87.86% (1556) who had been using the Internet for more than 5 years. During the survey period, 25.86% (458) were working as usual, 27.67% (490) were unemployed, 22.92% (406) were teleworking and 23.55% (417) were unemployed due to the pandemic and containment measures. Professional profiles are detailed in table 1. Regarding the population density, 7.4% (131) of participants were in a low (rural), 19% (335) in a medium (urban) and 73.7% (1305) in a high population density area.

**Table 1.** Characteristics of the included participants (N = 1771)

| Characteristics | % or mean (n or SD) |
| --- | --- |
| Gender |  |
| Female | 91.25 (1616) |
| Male | 7.96 (141) |
| Other | 0.79 (14) |
| Age (yrs) | 32.4 (13.39) |
| Smartphone/computer usage time (> 6 hrs/days) | 58.78 (1041) |
| Internet experience (> 5 yrs) | 87.86 (1556) |
| Profession |  |
| Farmer holding | 0.80 (14) |
| Artisan, dealer or business manager | 3.16 (55) |
| Manager or intellectual profession | 6.32 (112) |
| Intermediate profession | 5.59 (99) |
| Employee | 38.74 (689) |
| Worker | 4.18 (74) |
| Retired | 2.94 (52) |
| No professional activity | 38.18 (676) |

A total of 27.83% (493) declared they had symptoms of Covid-19, 21 participants tested positive, 11 negative and 461 were not tested. Of the 1771 respondents, 61.66% (1092) would have liked to be screened for Covid-19.

The four groups included 497 patients with asthma (from the “Vik Asthm” chatbot), 360 patients with breast cancer (from the “Vik Breast” chatbot), 459 patients with depressive disorder (from the “Vik Depression” chatbot) and 455 in the Vik Migrain group.

### 3.2 Findings

The global mean PDI score was 13.48 (8.02). Scores for each item are shown in table 2.

**Table 2.** Mean score, standard deviation and quartiles for each item of the PDI.

|  | Q1 | Q2 | Q3 | Q4 | Q5 | Q6 | Q7 | Q8 | Q9 | Q10 | Q11 | Q12 | Q13 | Total |
| --- | --- | --- | --- | --- | --- | --- | --- | --- | --- | --- | --- | --- | --- | --- |
| **Mean** | 1,05 | 1,52 | 1,53 | 1,51 | 0,29 | 0,79 | 2,44 | 1,04 | 0,89 | 1,23 | 0,67 | 0,1 | 0,41 | 13,48 |
| **std** | 1,15 | 1,29 | 1,3 | 1,35 | 0,77 | 1,17 | 1,26 | 1,24 | 1,16 | 1,23 | 1,07 | 0,43 | 0,85 | 8,02 |
| **25%** | 0 | 1 | 1 | 0 | 0 | 0 | 1 | 0 | 0 | 0 | 0 | 0 | 0 | 7 |
| **50%** | 1 | 1 | 1 | 1 | 0 | 0 | 2 | 1 | 0 | 1 | 0 | 0 | 0 | 12 |
| **75%** | 2 | 2 | 2 | 2 | 0 | 1 | 4 | 2 | 2 | 2 | 1 | 0 | 1 | 18 |

In total, 38.06% (674) of the respondents had psychological distress (score ≥15). The ANOVA analysis showed that sex (p=0.00132), unemployment (p=7.16×10-6) and depression (p=7.49×10-7) were significantly associated with a higher PDI score. Patients using their smartphone or computer more than one hour a day also had a higher PDI score (p=0.02588). There was no significant difference between groups (p>0.05; figure 1)

**Figure 1.**
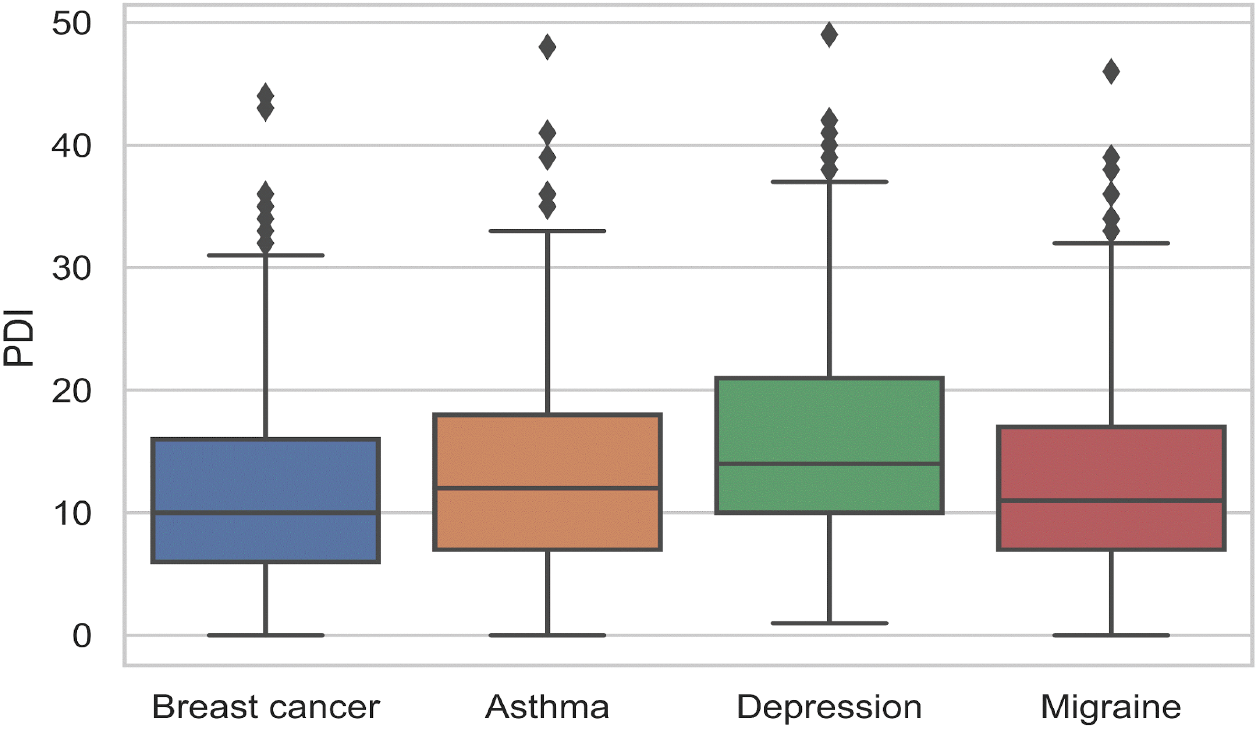
PDI for each group of participants.

Binomial logistic regression shows that patients with depression had a 53% (OR 1.53 [1.18-1.99]) increased risk (p=0.00134) of developing PD, while patients under 34 years old had a 72% (OR 1.72 [1.04, 2.89]) increased risk (p=0.03622), women a 63% (OR 1.63 [1.12-2.38] increased risk (p=0.01080) and unemployed patients a 38% OR 0.38 [1.12-1.71]) increased risk (p=0.00242). PDI was significantly higher in regions with higher Covid-19 prevalence (p=0.02355) (Pearson correlation coefficient = 0.58) (figure 2). The negative emotions factor was significantly associated with depression (p=2.71×10^-11^), age higher than 65 y.o. (p=0.02225), being a woman (p=0.00492), living in a low population density area (p=0.01330), being unemployed (p=1.38×10^-6^) and using a smartphone or computer more than 6 hours a day (p=0.03543). The life threat and bodily arousal factor was significantly associated with depression (p=2.95×10^-6^), age younger than 34 y.o. (p=0.01579), being a woman (p=0.00331), living in a low population density area (p=0.00593) and being unemployed (p=0.01077).

**Figure 2.**
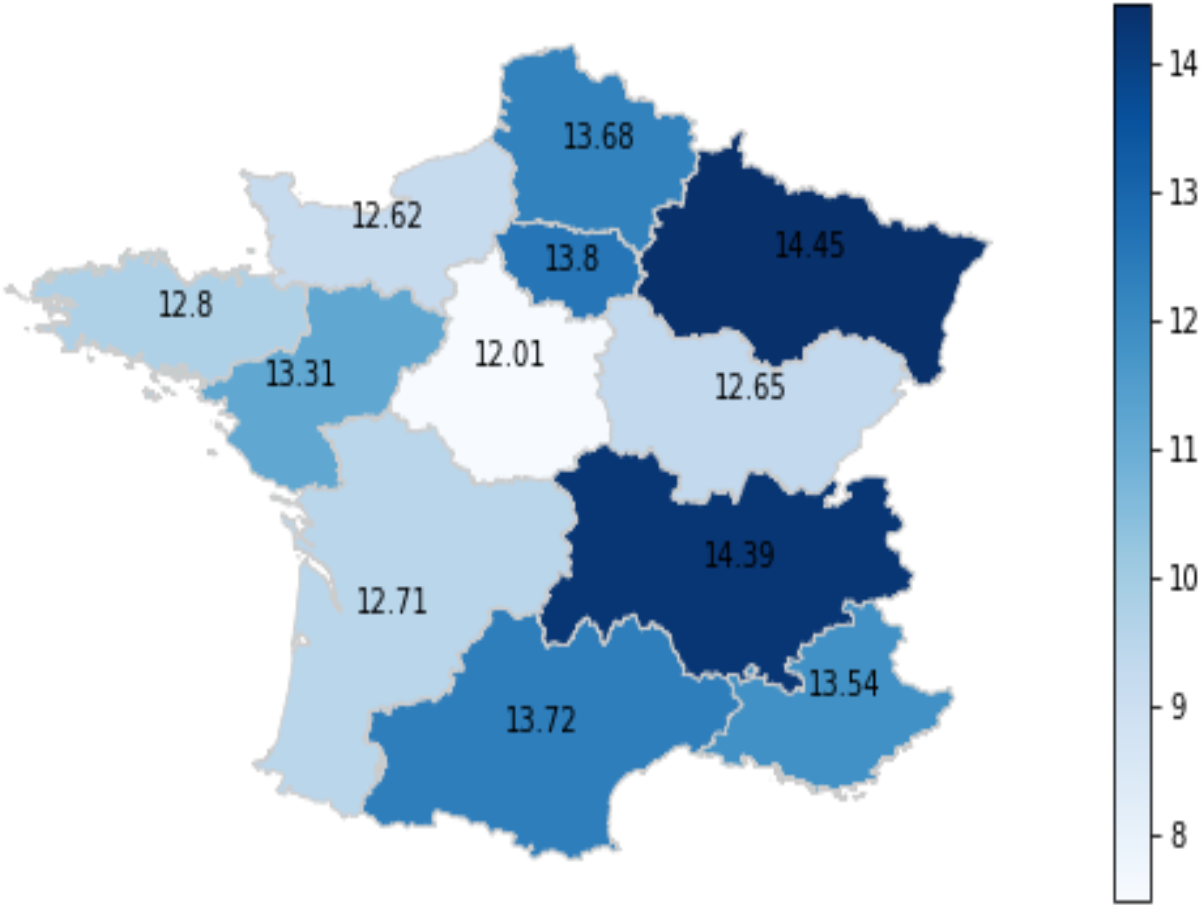
Average PDI in each of the 11 regions of France.

## 4. Discussion

The first case of COVID-19 was diagnosed in December 2019 in Wuhan, China and has since brought unprecedented efforts from governments all over the world to limit its spread. These steps have included social distancing and global shutdowns. Their precise consequences on mental health are still unknown. It is currently considered that the risk for mental health is outweighed by the need to prevent infections.

The available literature on the mental health consequences of pandemics are more focused on the sequelae of the infection, however other catastrophic events, such as the World Trade Center terrorist attacks, are followed by increase in depression and PTSD cases, substance abuse, domestic violence and child abuse (28). In that regard, the 2006 SARS epidemic also induced an increase in PD and PTSD in patients and clinicians (29). COVID-19 could also have the same effect, specifically because of the strong mitigation strategies that have been enforced all over the world, on a scale never seen before, but also because of the major economic disruptions it has induced (30).

The aim of our study was to quantify Psychological Distress and the risk of Post-Traumatic Stress Disorder on a national scale in a country that has been hard-hit by COVID-19, France. Using a chatbot-administered standardized and validated tool specifically designed to rate PD, the Psychological Distress Index (8). We showed that, in 4 groups of patients at-risk to develop PD and PTSD, the prevalence of psychological distress was high (38.06%, n=674). These patients are also at high risk to develop partial of full PTSD six-week after the evaluation, as shown by Bunnel et al. (9). Within those 4 groups of patients, women, unemployed (p=7.16×10^-6^) and depressed (p=7.49×10^-7^) patients had significantly higher PDI score. Interestingly, patients using their smartphone or computer more than one hour a day also had a higher PDI score (p=0.02588). This could also highlight the potential negative psychological impact of information and/or social networks in the context of such an event.

Our study is the first to assess PD in patients during a pandemic on a national scale. There are limitations that should be considered when interpreting our results: first, a majority of participants were women (91.25%). This is due in part to the fact that one of the 4 groups explored consisted in breast cancer patients, but it could also show that men are less likely to participate in this kind of online self-reported survey. This fact could potentially bias the results and specifically the value of the features we found to be associated with a PDI over 14 (predictive of PTSD). Another limitation is due to the sampling technique itself, relying on groups of patients already using the chatbots, excluding patients not using them. This study still holds interesting results because of the large cohort of respondents, the adequate geographical spread across France and the sampling time frame that corresponds to the pandemic peak in France.

Other studies have been conducted to measure the impact of COVID-19 among the general population. In Italy, Rossi et al conducted a web-based survey on 18,147. They found high rates of negative mental health outcomes three weeks into the COVID-19 lockdown: 37% of the participants declared they had symptoms of PTSD, 17.3% of depression and 20.8% of anxiety. Like in our study, the majority of respondents were women (79.6%).

Overall, policy makers are rightfully concerned by the potential negative effects on public health of COVID-19, beyond the pandemics itself. In the UK, psychological first aid guidance has been issued by Mental Health UK (31). In France, several psychological support hotlines have been created for healthcare professionals (32) and the general public (33). The precise mental health sequelae of the pandemic are still unknown but should not be neglected. In the coming weeks, months and years, we need to thoroughly investigate these consequences to be able to correctly address them. Specific efforts should be made to lower the risk of PD, depression, suicide, substance abuse and domestic violence, otherwise the long-term consequences of the COVID-19 pandemic could be even more dire, should they remain unexplored, unaddressed, and ultimately forgotten.

## Conclusion

COVID-19 has a significant impact on psychological distress in patients with breast cancer, asthma, depression and migraine: 38% of participants have a PDI equal or over 15. This population is also at increased risk of partial or full Post-Traumatic Stress Disorder. Specifically, women, unemployed and depressed patients are at an even higher risk. Patients using their smartphone or computer more than one hour a day are also at higher risk to develop PD. These measures call for systematic evaluation of the consequences of the COVID-19 pandemic in countries where lockdowns were enforced (2.5 billion people as of May, 3rd 2020).

## Data Availability

BC had full access to all of the data in the study and takes responsibility for the integrity of the data and the accuracy of the data analysis. The datasets used and analyzed during the current study are available from the corresponding author on reasonable request.

## Author contributions

Study concept (BC, JEB), drafting of the manuscript and supervision (BC, JEB, GD), acquisition of data (GD, AG), statistical analysis (AG), interpretation of data (BC, JEB), critical revision of the manuscript for important intellectual content (All authors).

